# Extreme weather events and dengue in Southeast Asia: a regionally-representative analysis of 291 locations from 1998 to 2021

**DOI:** 10.1101/2024.10.24.24316055

**Authors:** Sophearen Ith, Xerxes Seposo, Vitou Phy, Kraichat Tantrakarnapa, Geminn Louis C. Apostol, Pandji Wibawa Dhewantara, Rozita Hod, Mohd Rohaizat Hassan, Hidayatulfathi Othman, Mazrura Sahani, Jue Tao Lim, Ha Hong Nhung, Nguyen Hai Tuan, Ngu Duy Nghia, Taichiro Takemura, Inthavong Nouhak, Paul Lester Carlos Chua, Alex R Cook, Felipe J Colón-González, Masahiro Hashizume

## Abstract

**Background:** Climate change, leading to more frequent and intense extreme weather events (EWEs), could significantly impact dengue transmission. However, the associations between EWEs and dengue remains underexplored in the Southeast Asia (SEA) region. We investigated the association between selected EWEs (i.e. heatwaves, extremely wet, and drought conditions) and dengue in the SEA region.

**Methods and Findings:** Monthly dengue case reports were obtained from 291 locations across eight SEA countries between 1998 and 2021. Heatwaves are defined as the monthly total number of days where temperatures exceed the 95th percentile for at least two consecutive days. Droughts and extremely wet conditions are defined by a self-calibrating Palmer Drought Severity Index (scPDSI). We implemented a generalized additive mixed model coupled with a distributed lag non-linear model to estimate the association between each EWE and dengue. Months with fewer than 12 heatwave days increased dengue risk with delayed effect after two months lag, compared with months without any heatwave. Highest dengue risk is at 7 heatwave days (RR=1·28; 95%CI: 1·19,1·38). Compared to normal conditions (i.e. scPDSI=0), drought conditions (i.e. scPDSI=–4) were positively associated with dengue risk (RR=1·85; 95%CI: 1·73,1·99), while extremely wet conditions (i.e. scPDSI=4) have reduced dengue risk (RR=0·89; 95%CI: 0·87,0·91). Although the findings of this study are significant, its limitations arise from the inconsistency of dengue case reporting, which might complicate dengue risk estimation.

**Conclusions:** This study shows that the delayed effect of heatwaves and drought conditions magnifies the risk of dengue in the SEA region. Our findings offer stakeholders sizeable amount of time to organize and implement public health interventions in minimizing the prospective dengue risk, posed by EWEs in the context of climate change in SEA. Future research may focus on factors associated with dengue risk variations within SEA region to facilitate the development of location-based, tailor-fit mitigation and preventative interventions.

## Introduction

Dengue is caused for Flaviviruses, transmitted by the infectious bite of *Aedes aegyti,* and *Ae. albopictus* mosquitoes. Dengue poses an alarming impact on human health, and the global economy, particularly in the Southeast Asian (SEA) region [1]. The climate in the SEA region, characterized by persistent hot and humid weather throughout most of the year, offers optimal environmental conditions for mosquito development and dengue transmission [2]. According to a recent projection study, dengue incidence in SEA is expected to reach its peak in the 21st century [3]. By the end of this century, the length of dengue transmission season in the SEA region is expected to be longer with a rise in number of population at risk of up to 696 million additional people [4].

Climate change, including extreme variations in precipitation and prolonged extreme temperatures, could modify the population density of the dengue vector [2]. Consequently, variation in the dengue transmission risk may occur following Extreme Weather Events (EWEs), including heatwaves, drought, and flood [5–7]. Briefly, heatwaves, defined as prolonged periods of extreme heat over a given threshold, could result in higher mosquito populations as heatwaves can accelerate the early development stage of mosquitoes life cycle [8]. Drought conditions have been also linked to increased dengue transmission risk via water storage in man-made containers [9]. Moreover, while cumulative precipitation might enhance mosquito abundance, heavy precipitation flushes away breeding sites, thereby decreasing mosquito populations [10].

In the context of a warming climate, EWEs are anticipated to become more frequent and intense [11]. Predictive models suggest that the dengue-endemic SEA region is becoming more susceptible to the adverse effects of climate change that included but are not limited to heat stress, heavy precipitation, widespread flooding both inland and in coastal zones, prolonged dry spells and acute deficits in water supply [11]. Heatwave intensity is projected to rise by 0·5 to 1·5 °C above a given global warming threshold, with an increase in duration of 2 to 10 days per 1°C of global temperature rise [12]. Climate models under Representative Concentration Pathway (RCP4·5) scenario and the 14-model ensemble mean from Coupled Model Intercomparison Project Phase 5 (CMIP5) projected an increasing trend of drought in SEA in the future [13].

Despite the substantial growth in evidence regarding the impacts of EWEs on human health, their effect on mosquito-borne diseases like dengue is limited especially in SEA, where dengue is endemic [14]. In line with WHO’s Global Vector Control Response operational framework, it is essential to investigate the impact of climate factors, particularly EWEs, on dengue using a multi-country approach in generating evidence required to efficiently prevent and manage dengue transmission and respond to subsequent outbreaks [15]. To our knowledge, this is the first study investigating the associations between multiple EWEs (i.e. heatwaves, extremely wet and drought conditions) and dengue in the SEA region, utilizing a robust statistical modelling approach.

## Methods

### Data collection Dengue data

Monthly dengue count data were obtained from the Southeast Asia Research on Climate change and Dengue (SEARCD) collaboration platform for 268 locations in seven countries including Indonesia (33 provinces during 2010–2020), Lao PDR (2 provinces during 2005–2021), Malaysia (14 states and federal territories during 2010–2017), the Philippines (79 provinces during 2010–2020), Singapore (1 city state during 2010–2017), Thailand (76 provinces during 2003–2021), and Vietnam (63 provinces during 2011–2021). Briefly, SEARCD is a platform with collaborators from the seven SEA countries that encourages research on meteorological factors and dengue. Additionally, monthly dengue count data from 23 locations in Cambodia during 1998–2010 was obtained from project Tycho 2·0 [16].

### Temperature and precipitation

Monthly meteorological data, including mean temperature, mean dew point temperature, and total precipitation, were obtained from the ERA5-Land dataset generated by the European Centre for Medium-Range Weather Forecasts (ECMWF) (appendix pp. 2) [17]. ERA5-Land is a reanalysis data set providing climate information at a fine spatiotemporal resolution (0·1°×0·1° grid) for the whole global land surface [17].

### Heatwave definitions

Heatwaves were defined using relative thresholds, which consider the long-term daily mean temperature at each location. Heatwave were defined as two or more consecutive days with a daily mean temperature surpassing the 95^th^ percentile of its distribution in each specific location [18,19]. Hourly temperature data were averaged to compute the daily mean temperature for each location. The total number of heatwave days (HWt) was then calculated for each month and location. If a heatwave begins in the final days of one month and continues into the next, the heatwave days are included to each respective month (Table S2).

### Wet and drought conditions

We used the Palmer Drought Severity Index (PDSI) to assess the effect of drought and extremely wet conditions on dengue. PDSI is a widely recognized standardized index for tracking drought and long-term variations in aridity and is computed by considering soil moisture content, the projected rate of evapotranspiration (i.e. the evaporation from soil under adequate water availability, considering mean daily temperature and numbers of days in the month) and rainfall amounts [20]. Here, we used the self-calibrating PDSI version (scPDSI) since it represents a geographically comparable index by calibrating a distinct normal condition for each location [21]. The scPDSI data were sourced at a geographical resolution of 0·5°×0·5° grid from the Climatic Research Unit gridded Time Series (version 4·05) corresponding to the specific time in each location [22]. The scPDSI ranges from –10 (i.e., very dry conditions) to 10 (i.e. very wet conditions). Values below –4 or over 4 are categorized as drought and extremely wet conditions, respectively [21].

### Population data

Total population count for each location were obtained from the Gridded Population of the World Version 4 developed by the Socioeconomic Data and Application Center for the period 2000 to 2020 at five-year intervals [23]. Linear interpolation was used to estimate yearly total population values for each location over the study period.

### Statistical analysis

We specified a generalized additive mixed model (GAMM) coupled with a distributed non-linear model (DLNM) to investigate the association between EWEs and dengue [24,25]. A quasi-Poisson distribution was assumed to account for potential over-dispersion in the dengue data. The logarithm of the total population for each location was included as an offset to adjust dengue case count by population. A smoothing spline with 5 degrees of freedom (df) per year was included to control for potential seasonality (Table S4). A cross-basis function was included in each model using natural cubic spline with two equally spaced knots for heatwave and 3 df for scPDSI (i.e. the exposure dimension) and 3 df for the lag dimension based on exploratory analyses (Table S5). We included a smoothing spline with 3 df to control for potential nonlinear effects of monthly mean temperature and total precipitation based on previous study [26]. Long-term trends and interannual variability were accounted for incorporating an indicator term for each year in the time series. Unmeasured factors, such as public health interventions, were specified as unstructured random effects for each location. The reference value of heatwave was set at 0 heatwave days per month and scPDSI at 0. Models were fitted in R (version 4·3·1), using the packages *mgcv* 1·9·1 and *dlnm* 2·4·7 [24,25].

The algebraic definition of the EWE-dengue model is given by:

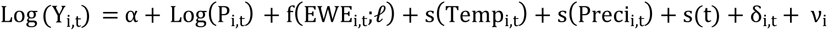

where Y_i,t_ indicates monthly dengue case at time t in location i, assumed to follow an over-dispersed quasi- Poisson distribution; α represents the intercept; Log(P_i,t_) represents the logarithm of the total population per location i at the time t, included as an offset; f(EWE_i,t_;*ℓ*) designate the cross-basis functions of either heatwave (HW) or scPDSI for maximum lag (*ℓ*) of 4 months; s(Temp_i,t_) and s(Preci_i,t_) denotes the smooth functions of temperature and precipitation, respectively; *s*(*t*) denotes the smooth seasonals trend of time; *δ*_*i,T*_ indicates long- term trend using indicator variables for each year T in each location i; and ν_i_ expresses unstructured random effects for each location.

### Sensitivity analysis

Multiple sensitivity analyses were carried out to assess the robustness of the results. We first examined the optimal exposure-response relationship in the cross-basis function by varying the df and placement of the knots, using natural cubic spline with two equally space internal knot, two knots at 25^th^ and 75^th^ percentile, and one knot at 50^th^ percentile. We further applied a varying function of seasonality including smoothing spline with 3df, 4df, cyclic cubic spline, and one pair of Fourier term. Maximum lags were also varied by 3 months, 5 months, and 6 months, respectively. We repeated the analysis with adjustment relative humidity and population density in the model to account for additional confounding effects. Analyses using the data from 1998 to 2019 were also conducted to determine the potential impact of the COVID-19 pandemic on our results. Various heatwave definitions were investigated including: (i) four or more days exceeding the 95^th^ percentile, (ii) two or more days exceeding the 97^th^ percentile, (iii) two or more days exceeding the 99^th^ percentile, (iv) four or more days exceeding the 97^th^ percentile, and (v) four or more days exceeding the 99^th^ percentile.

## Results

### Descriptive statistics

Table 1 shows the summary statistics of dengue cases and meteorological factors in each country. A total of 7,016,624 dengue cases was included in this study. The Philippines reported the highest mean annual dengue cases (196,719 cases), followed by Vietnam (121,822 cases) and Indonesia (112,156 cases). The monthly mean temperature, monthly total precipitation, mean annual heatwave days, and monthly scPDSI index at the country level ranged between 24·1°C and 26·8°C, 138mm and 254mm, 14·2 days and 17·2 days, and –0·3 and 0·3, respectively. State of Selangor in Malaysia showed the highest monthly dengue incidence per 100,000 people followed by Khah Hoa in Vietnam (Figures 1A). The lower mean annual number of heatwave days was observed in south-eastern locations (Figures 1B). Eastern Philippines and Northern of Vietnam and Cambodia indicate wetter conditions (Figures 1C). Additional descriptive results of each climate variable and dengue incidence for each location are shown in Table S3.

**Figure 1:**
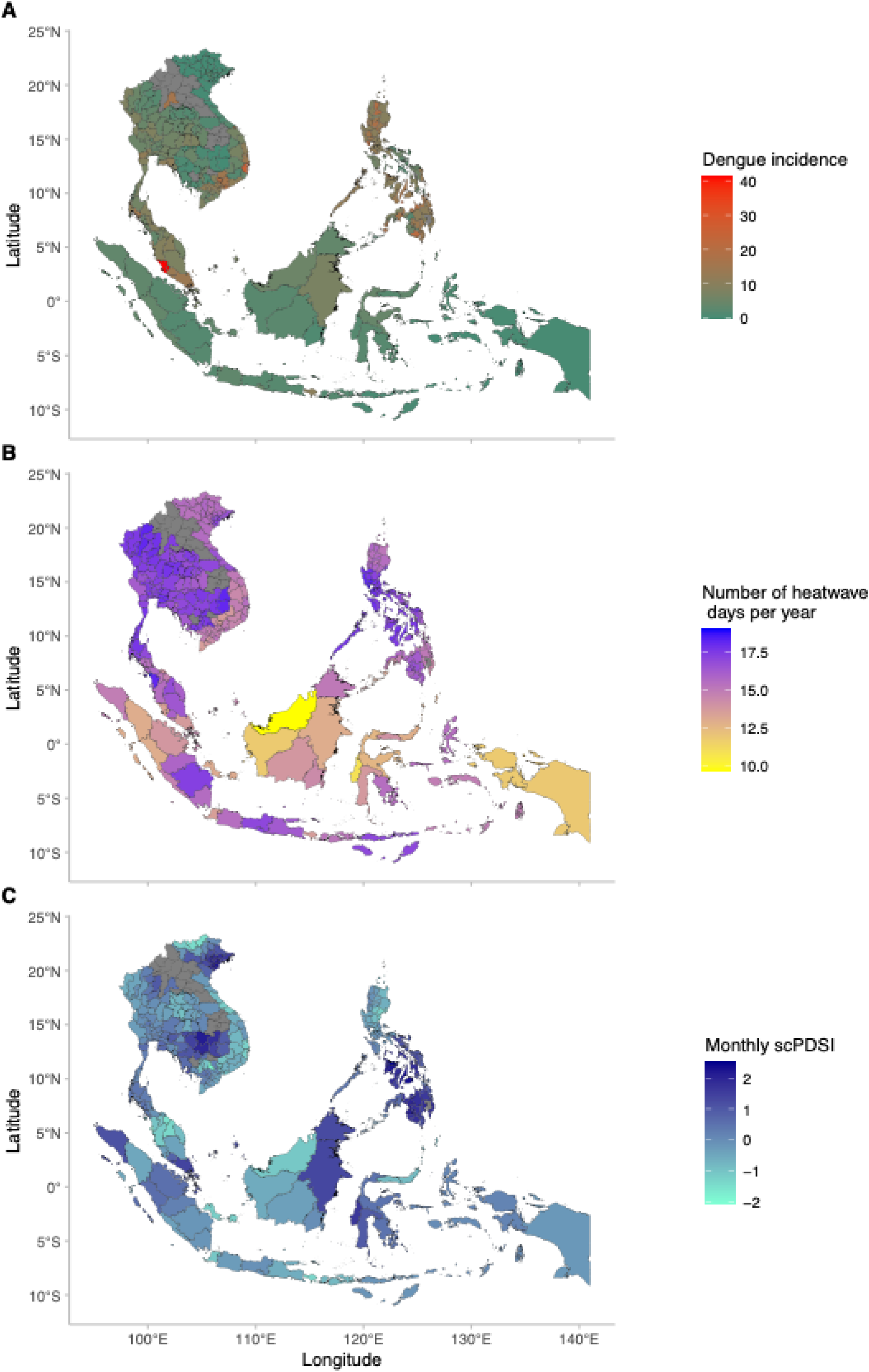
Map of dengue case, heatwave, and scPDSI in each location of this study. (A) Median monthly dengue incidence per 100,000 population in each location. (B) Mean annual number 05 of heatwave days in each location. (C) Median monthly scPDSI index in each location. Grey areas have 06 no data. scPDSI stands for self-calibrated Palmer drought severity index.

**Table 1:** Summary statistics of dengue cases, monthly temperature, total precipitation, heatwave days and scPDSI index in eight countries.

|  | Period | No of locations | Total dengue case | Mean annual dengue case | Monthly mean temperature<sup>a</sup><br>[mean (SD)] | Monthly total precipitation<sup>a</sup><br>[mean (SD)] | Annual number of heatwave days<sup>a,b</sup><br>[mean (SD)] | Monthly scPDSI<sup>a</sup><br>[mean (SD)] |
| --- | --- | --- | --- | --- | --- | --- | --- | --- |
| Cambodia | 1998–2010 | 23 | 152,058 | 11,697 | 26·8 (0·9) | 151 (37) | 17·2 (2·4) | 0·3 (0·8) |
| Indonesia | 2010–2020 | 33 | 1,233,718 | 112,156 | 24·7 (1·2) | 254 (51) | 14·2 (4) | 0·1 (0·6) |
| Lao PDR | 2005–2021 | 2 | 63,391 | 3,729 | 25·1 (0·6) | 158 (14) | 17·1 (2·6) | 0·1 (0·6) |
| Malaysia | 2010–2017 | 14 | 545,479 | 68,185 | 25·5 (0·8) | 222 (31) | 15·3 (5·9) | −0·3 (0·7) |
| Philippines | 2010–2020 | 79 | 2,163,905 | 196,719 | 25·1 (1·4) | 230 (36) | 16·5 (4) | 0·8 (0·9) |
| Singapore | 2012–2019 | 1 | 91,463 | 11,433 | 26·9 (0·6) | 207 (89) | 14·6 (2·9) | 0·8 (2·0) |
| Thailand | 2003–2021 | 76 | 1,426,568 | 75,083 | 26·1 (1·2) | 138 (32) | 17 (2·3) | 0 (0·4) |
| Vietnam | 2011–2021 | 63 | 1,340,042 | 121,822 | 24·1 (2·3) | 173 (26) | 15·2 (2·9) | 0 (0·8) |
|  | Total | 291 | 7,016,624 |  |  |  |  |  |

scPDSI stands for self-calibrated Palmer drought severity index; SD is standard deviation.
<sup>a</sup>: Location specific mean climate variable (SD)
<sup>b</sup>: 95<sup>th</sup> percentile for 2 consecutive days

### Heatwave-dengue association

Figure 2A presents the pooled cumulative (lag 0 to 4 months) association between the total number of heatwave days per month and dengue relative to month without any heatwave day. The association between the monthly number of heatwave days and dengue was non-linear and showed a significant positive association for months with less than 12 heatwave days and a negative association between months with 12 and 27 heatwave days. The dengue risk leveled off at months with 7 heatwave days (RR=1·28; 95%CI: 1·19,1·38) and reached the minimum dengue risk at month with 21 heatwave days (RR=0·49; 95%CI: 0·42, 0·57). The lag-response association of month with 7 and 21 heatwave days show dengue risk increase after lag 1 and 3 months, respectively (Figure 2B). The lag-response association showed a negative association across all numbers of heatwave days per month within a lag of two months (Figure 2C). However, the dengue risk increases mostly after the lag of two months.

**Figure 2:**
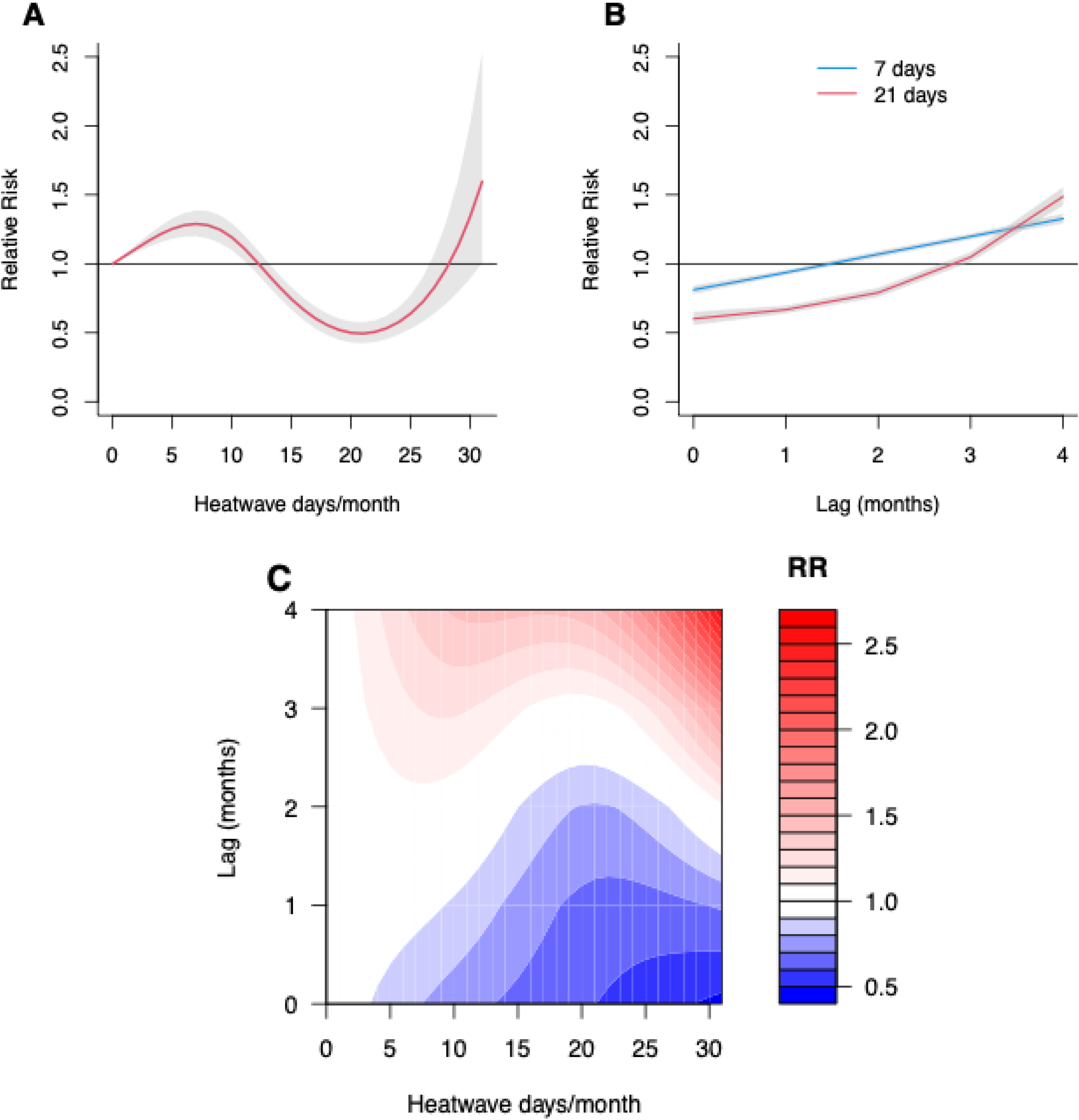
Association between the number of heatwave days per month and dengue incidence in 291 locations across the eight countries, relative to month with no heatwaves. (A) Cumulative exposure–response associations (with 95%CI, shaded grey), (B) lag-response association curves of specific monthly number of heatwave days (with 95% CI, shaded grey), (C) Contour plot of the association between monthly number of heatwave days and dengue. The deeper the shade of red, the greater the increase in the RR of dengue, and the deeper the shade of blue, the greater the decrease in the RR of dengue. CI indicates the confidence intervals; HW stands for number of heatwave days per month; RR is the relative risk.

### scPDSI-dengue association

Figure 3A shows the overall cumulative (lag 0 to 4 months) association between monthly scPDSI and dengue, compared to a value zero of scPDSI. The association between monthly scPDSI and dengue was non-linear and followed a reverse J-shaped curve association with minor upward tail and the minimum dengue risk observed at 2·5 scPDSI value (RR=0·89; 95%CI: 0·87,0·91). Drought conditions (i.e. scPDSI values equal to or lower than –4) show a significant positive association with dengue incidence (RR=1·85; 95%CI: 1·73,1·99). In contrast, the extremely wet conditions show a significant negative association with dengue risk (RR=0·92; 95%CI: 0·87,0·96) at scPDSI value between 4 and 5·8. An increase in dengue risk is observed when scPDSI is greater than 5·8, though the result is not significant.

**Figure 3:**
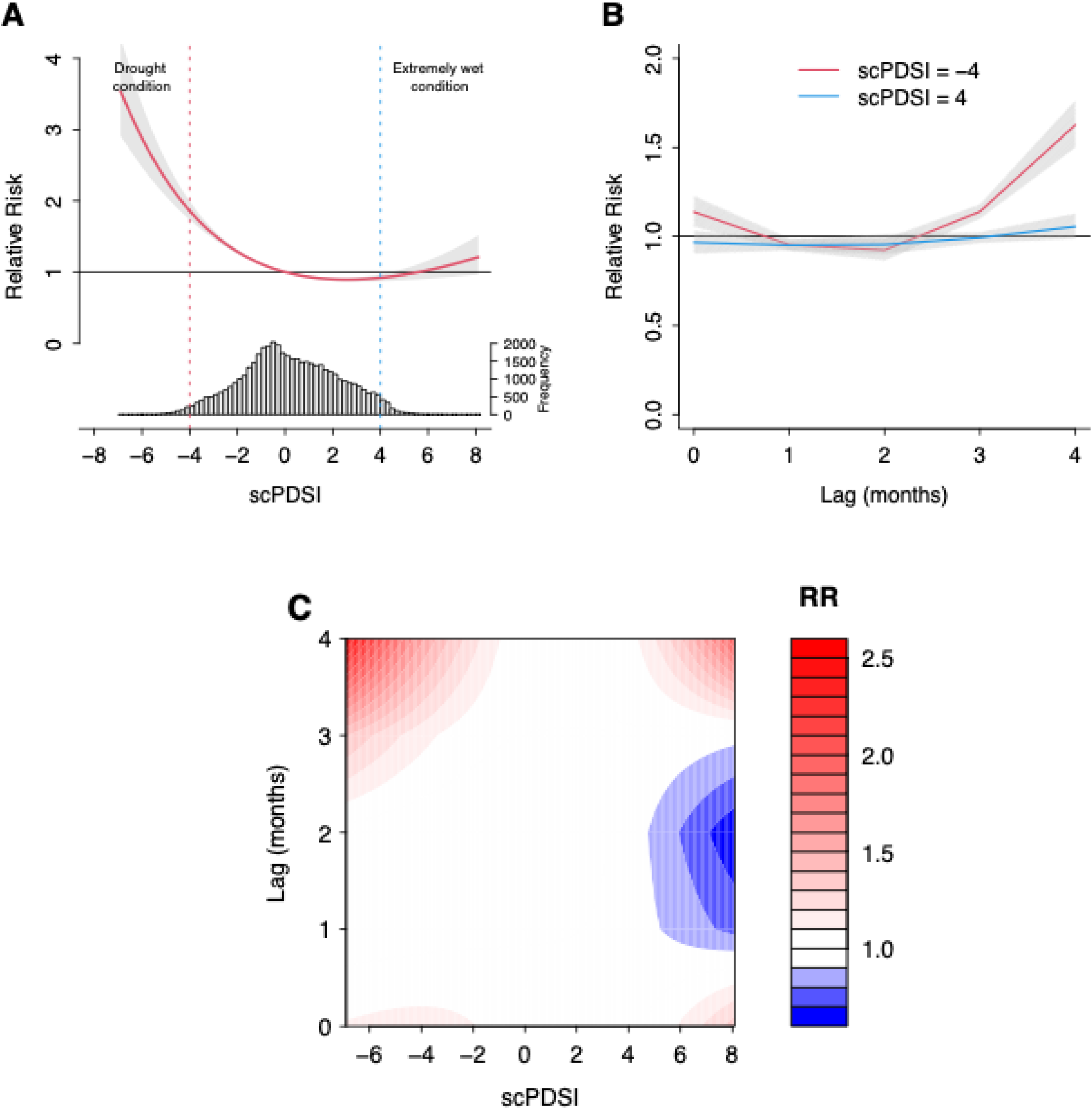
Association between scPDSI and dengue incidence in 291 locations across the eight countries, relative to a scPDSI value of zero. (A) Cumulative exposure–response associations (with 95%CI, shaded grey), with related PDSI distribution. Two vertical lines indicate the cut-off value for drought as a dotted line in red, and cut-off blue for extremely wet conditions as a dash-dotted line in blue. (B) lag-response association curves of specific scPDSI (with 95%CI, shaded grey). (C) Contour plot of the association between scPDSI and dengue. The deeper the shade of red, the greater the increase in the RR of dengue, and the deeper the shade of blue, the greater the decrease in the RR of dengue. CI indicates the confidence intervals; RR is the relative risk; scPDSI stands for self-calibrated Palmer drought severity index

The lag-response association of drought conditions shows both immediate and delayed positive association on dengue risk within lag 0 and after two months lag (Figure 3B). The lag pattern of extremely wet conditions shows a positive effect three months lag, suggesting a delayed positive effect on dengue risk (Figure 3B). When the scPDSI exceeds 5·8, there is an increased relative risk observed at a 1 month and after a 3-month lag (Figure 3C).

### Sensitivity analysis

The pooled effect estimates for each EWE on dengue incidence were generally robust, amidst varying adjustments for seasonality, maximum lag, as well as inclusion of relative humidity and population density (Figures S8, S9, S10). Changing the df for the cross-basis function did not change the direction nor the intensity of the association between scPDSI and dengue but did change it for heatwaves (Figure S7). For the heatwave model, result shows a reduction in dengue relative risk regardless of the monthly number of heatwave days when two internal knots were used at the 25^th^ and 75^th^ percentile or one internal knot at the 50^th^ percentile (Figure S7). Analyses using the data from 1998 to 2019 shows no impact of the COVID-19 pandemic on our results (Figure S11).

Using various heatwave definitions in the sensitivity analysis did not change the direction of the association, except for heatwave with the 99^th^ percentile for 2 and 4 consecutive days (Figure S12). With the definition of 99^th^ percentile for 2 and 4 consecutive days, there was a negative association between zero to 16 heatwave days per month and dengue. This was likely because of significantly fewer counts of months with heatwave days when the strictest heatwave definition was used.

## Discussion

We examined the associations between several EWEs including heatwave, drought, and extremely wet conditions on dengue incidence in the SEA region. Months with fewer than 12 heatwave days showed increased dengue risks with a delayed effect observed after two months lag. This may partially be explained by the role of temperature on vector population since mosquitoes can survive even under continuous exposure to extreme heat amidst these heatwave durations [8]. In addition, heatwave event potentially facilitates vector population growth in the early development phase leading to an increased dengue transmission in the latter phase [8]. Also, the number of heatwave days may associated with a faster rate of viral replication within the vector and with a shorter extrinsic incubation period— the time required for dengue virus to become transmissible to another host after initial infection of a mosquito [2]. However, during the intense duration of heatwaves, adult mosquito survival rates and feeding activities start to decline, explaining our result of a reduction in dengue risk at month with more than 12 heatwave days [2].

Our lag-exposure association result showed heatwaves more likely to increase dengue risk after a lag of two months. Comparing this finding with previous studies has its own limitations due to differences in heatwave definitions and statistical approaches used. However, a study in Hanoi, Vietnam, showed that heatwaves have an overall inhibitive effect at a shorter lag but were associated with increased magnitude of outbreaks at later lags [5]. In China, similar findings whereby heatwaves increase the risk of dengue outbreaks after six weeks were also observed [18]. This could be attributed to diapause behavior in mosquitoes, which is the inactive state where the dengue vectors halt their development and hatch to withstand harsh climatic conditions such as extreme heat or dry conditions [27]. The subsequent drops in temperature and increased humidity create favorable conditions for the continuation of the reproduction cycle of dengue vectors, ultimately leading to an increase in the dengue vector population [28]. The lags in mosquito outbreaks after a heatwave could result from the influence of climatic fluctuations on mosquito biology, serving as an adaptation mechanism to cope with unfavorable climate conditions [28].

Additionally, the association may be related to the change in human activities during and after heatwaves. During the heatwave, people tend to stay indoors in air conditioned environment, which limits vector-host contact [5]. However, lower temperatures after heatwaves encourage people to spend more time outside, increasing the likelihood of being bitten by mosquitoes. Another explanation might be an impairment of the host immunological response after exposure to certain sustained high temperature during heatwave [29]. Prolonged heat stress can reduce the ability of human body to mount an effective immune response, potentially making it easier for the dengue virus to be viable as an infection [29].

scPDSI index and dengue has a non-linear association with drought conditions showing an increase in dengue risk and extremely wet conditions showing a reduction in dengue risk. The similar association was reported in other studies in Brazil and China [7,30]. We noted that drought was positively associated with dengue risk at lags 0 and two months. This is likely because severe dry conditions increased water storage in artificial containers, contributing to an increase in reproduction sites and the proliferation of mosquitoes [9]. Changes in water storage behavior may enhance the abundance of aquatic habitat for mosquitoes, given that their eggs can survive for up to 120 days in a drought environment [9]. The lag observed between a period of extreme drought and an uptick in dengue risk may be attributable to the incremental changes in domestic water storage habits as households adapt to the scarcity of water. This change might prompt individuals to adopt measures such as storing water in makeshift containers around the home during periods of water shortage. This finding indicates the importance of ongoing monitoring of drought conditions and the prompt implementation of emergency measures to reduce preventable negative effects of dry conditions on dengue risk.

We found that extremely wet conditions showed a reduction in dengue risk, which is in line with a study in Singapore, where a significant reduction in dengue outbreak risk followed the flushing effect of precipitation [10]. Non-standing water could flush away the mosquito eggs and shelter, consequently decreasing the dengue risk [6]. However, our results showed that severely wet conditions increase dengue risk, which could potentially be due to stagnant waters amidst severe wet conditions favoring development and reproduction of mosquitoes [6]. Moreover, severely wet conditions may also contribute to hydrological natural disasters, including floods or tropical cyclones, leading to significant deterioration of infrastructure or electricity, increasing the risk of close contact between dengue vectors and humans [6]. However, the impact of severely wet conditions needs further investigation to understand this phenomenon.

## Strength and limitations

This research possesses a few strengths. First, this is the first study investigating of the EWEs-dengue association in the SEA region. This study was carried out in numerous subnational locations across most of the SEA region, representing different ranges of exposure levels and socio-demographic characteristics. By using a pooled design approach, this study provided strong evidence of the relationship between EWEs and dengue at the regional scale. Second, We used GAMM and DLNM that captures the complex non-linear and delayed relationships between climate variables and dengue incidence [24,25]. The use of GAMM enables the consideration of both spatial and temporal variations, which means the model can account for differences across locations and over time—factors that are critical when studying a vector-borne disease like dengue [24].

However, some limitations need to be acknowledged. First, a significant limitation is the use of a single set of modeling parameters across all the locations, which may heavily impact location/country-specific estimations. Specifically, the chosen modeling may not fit the data well in certain locations, even if it results in the best overall fit. However, the robustness of the results to various modeling choices and selections was assessed by exploring different model specifications for exposure and lag. Additionally, dengue case reporting can vary due to regional and temporal differences in diagnosis approaches, leading to over- or under-reporting of dengue cases (Table S1). These variations may complicate accurate assessments of dengue risk, potentially skewing understanding of the climate true impact on dengue. Moreover, utilizing monthly data to study dengue risks may not capture short-term climate effects and dengue incubation periods. Also, relying on monthly total heatwave days might overlook the nuances of individual heatwave episode impacts on dengue transmission. The potential influence of serotype information on the climate-dengue association is acknowledged. Due to data availability limitations, the analysis could not be stratified by dengue serotypes, highlighting the need for future research in this aspect.

## Conclusions

Given the anticipated rise in the number and intensity of EWEs, this research offers insights into estimating the dengue risk associated with heatwave, drought, and extremely wet conditions introduced by the global climate in a real-world setting. Our study showed that delayed heatwaves and drought effects contribute to increased dengue risk. These findings provide health managers and policymakers a better understanding of the contribution and importance of EWEs on dengue risks, and likewise serves as a groundwork in developing parallel strategies and interventions targeting both EWE adaptation and dengue response suited to the regional SEA context.

## Acknowledgements

We would like to extend our sincere gratitude to Dr. Futoshi Hasebe and all collaborators for their valuable contributions to this study. We also thank Japan Agency for Medical Research and Development (AMED) (JP20wm0125006) for their support.

## Author Contributions

IS, XS, and MH conceptualized the study. XS collected and compiled dengue data. IS collected the climate and population data, ran the modelling, and drafted the manuscript. VP provided the code for downloading climate data. MH, XS, FJC, and ARC guided the methods, presentation of the result and discussions. XS and MH verified the modeling outputs. IS and XS had full access to all the data in the study. All authors reviewed, approved the manuscript, and had final responsibility for the decision to submit for publication.

## Data sharing

The dengue case data, except for Cambodia was obtained from collaboration platform of the Southeast Asia Research on Climate Change (SEARCD) under a data agreement and cannot be made publicly available. Dengue data of Cambodia can be freely downloaded from Project Tycho (https://doi.org/10.25337/T7/ptycho.v2.0/KH.38362002). Climate data can be downloaded from Copernicus (https://doi.org/10.24381/cds.68d2bb30). The self-calibrating PDSI can be sourced from the Climate Research Unit (https://crudata.uea.ac.uk/cru/data/hrg/). Population data can be downloaded from the Socioeconomic Data and Application Center (https://doi.org/10.7927/H4PN93PB). Researcher can refer to the corresponding authors for information on accessing the data. Code sources are available from the corresponding author upon reasonable request.

